# Advantages to interspace-level analyses over person-level analyses when studying relationships between lumbar spinal degeneration findings : a cross-sectional study

**DOI:** 10.1101/2024.12.25.24319627

**Authors:** Pradeep Suri, Elizaveta E. Elgaeva, Jeffrey G. Jarvik, Sean D. Rundell, Yakov A. Tsepilov, Frances M.K. Williams, Patrick J. Heagerty

## Abstract

**Purpose:** To examine associations between lumbar intervertebral disc degeneration (LDD) and type II Modic changes (MC) when retaining information at each interspace (“interspace-level analysis”), as compared to aggregating information across interspaces as is typically done in spine research (“person-level analysis”). The study compared results from (1) interspace-level analyses assuming a common relationship across interspaces (the “interspace-level, common-relationship” approach), (2) interspace-level analyses allowing for interspace-specific associations (an “interspace-level, interspace-specific” approach), and (3) a conventional person-level analytic approach.

**Methods:** Adults in primary care (n=147) received lumbar spine magnetic resonance imaging (MRI) and neuroradiologist-evaluated assessments of prevalent disc height narrowing (DHN), type II MC, and other LDD parameters. Analyses examined associations between DHN and type II MC in interspace-level, common-relationship analyses, interspace-level, interspace-specific analyses, and conventional person-level analyses.

**Results:** Cross-sectional, interspace-level, common-relationship analyses found large-magnitude DHN-type II MC associations (adjusted OR [aOR]=6.5, 95% confidence intervals (CIs) 3.3-13; p<0.001). The magnitude of this association was larger and more precise than that yielded by person-level analyses (aOR=2.9 [95% CI 1.2-7]), and substantially more precise than interspace-level, interspace-specific analyses which allowed the association between DHN and type II MC to vary across levels. Across exploratory analyses of disc signal intensity and other MC types, interspace-level, common-relationship analyses produced larger-magnitude and more precise associations than person-level analyses in most situations, and were more precise than interspace-level, interspace-specific analyses.

**Conclusions:** Interspace-level analytic approaches offer some advantages to person-level analyses that may be useful in understanding relationships between spinal degeneration findings.

## INTRODUCTION

An active area of spine research is the study of how lumbar spinal degeneration findings are related to each other. Analyses of relationships between different imaging-detected lumbar intervertebral disc degeneration (LDD) findings commonly take a “person-level” approach, with the unit of analysis being the individual person. Person-level analyses aggregate findings across spinal levels, resulting in coarse summaries of an imaging finding at any lumbar spinal level or across the lumbar spinal levels.^1-7^ However, spinal levels (or “interspaces”) are not independent from one another, due to their interrelated biomechanics and proximity to each another. Many complexities of relationships between interspaces can be avoided by person-level analytic approaches, which ignore interspace-level interactions and by doing so assume that such interactions are not important. Yet while person-level analyses of LDD are easy to conduct, they have potential problems. First, person-level analyses of LDD lose information by summarizing the information contained in the 5 lumbar interspaces, which may differ from each other with regards to typical LDD prevalence, biomechanics, and biology. This loss of information can lead to decreased precision and effect sizes. Second, person-level analytic approaches to spine analyses do not provide a complete framework that allows modelling of complex interspace-specific interactions that may exist between different LDD findings and combinations of findings. Therefore, in some situations, a person-level approach may impede the discovery of more complex interactions between spinal levels, degenerative findings, alignment, and other factors.

“Interspace-level” analytic approaches are an alternative to or complement to person-level analytic approaches that may offer advantages in some scenarios when studying relationships between LDD findings. Interspace-level analyses use each spinal *interspace* as the unit of analysis, rather than the *person*. Historically, two simple yet flawed approaches to multivariable interspace-level analyses have been widely used. One such approach treats each interspace as the unit of analysis but assumes that different interspaces within a person are independent.^4,8,9^ This assumption is clearly not true, given the interrelatedness of spinal levels within a person that exists due to biomechanics and biology, and can potentially affect estimates of precision.^10^ A second approach is to conduct multiple person-level analyses of LDD, each restricted to a specific interspace (e.g., L5-S1)^1,11-14^ or region (e.g., L4-S1).^5^ This approach requires multiple statistical comparisons and consequently can increase type I error. Neither of these simple approaches allow for explicit statistical tests of more complicated questions such as whether different imaging findings have significantly different relationships at different interspaces or regions. Moreover, neither approach mirrors the interrelatedness of spinal levels within a person that exists due to biomechanics and biology. Contemporary analytic methods such as generalized estimating equations (GEE)^15^ allow better ways to conduct interspace-level analyses, such as (1) interspace-level analytic approaches that assume a common relationship across interspaces (in which the unit of analysis is the spinal interspace rather than the person, which we refer to here as the “*interspace-level, common-relationship*” approach) and (2) interspace-level analytic approaches that allow for associations between different imaging findings to vary between different spinal interspaces (which we refer to here as the “*interspace-level, interspace-specific*” approach). Fewer spinal imaging studies have used interspace-level, common-relationship^16^ and interspace-level, interspace-specific approaches^17^ that account for the interrelatedness of spinal interspaces and their lack of independence.

In the current work, we compare and contrast the results of interspace-level vs. person-level analyses of spinal degeneration findings using an illustrative example of the relationship between two LDD findings, disc height narrowing [DHN] and disc signal intensity loss [DSI], and vertebral endplate or “Modic” changes (MC).^3,5^ LDD and MCs are generally associated in cross-sectional imaging studies, with varying magnitudes of association between studies.^3,5,11,16^ Given the low prevalence of type I and type III MCs,^11^ which limit power to detect associations with MCs, the current study focused on type II MCs.

The first study aim was to examine associations between LDD and type II MCs in a cross-sectional interspace-level analysis that assumes a common relationship across interspaces (the *interspace-level, common-relationship* approach). We hypothesized that LDD and type II MC would be significantly associated when using this approach. The second aim was to descriptively compare results from analyses of LDD and type II MC using different analytical approaches: (1) an *interspace-level, common-relationship approach*, (2) an *interspace-level, interspace-specific approach* that allows for interspace-specific associations, and (3) a conventional *person-level approach*. We hypothesized that the *interspace-level, interspace-specific approach* would be significantly different from the interspace-level, common-relationship approach.

## METHODS

### Overview

This was a secondary analysis of the Longitudinal Assessment of Imaging and Disability of the Back (LAIDBACK) study.^18,19^ LAIDBACK was a longitudinal cohort study of 147 adults without LBP at baseline who were followed for incident clinical symptoms such as LBP. Participants received baseline lumbar spine magnetic resonance imaging scans (MRIs) and repeat MRIs at 3-year follow-up. The analysis plan for the current work was completely pre-specified and publicly posted prior to any analyses.^20^ DHN was prioritized as the main LDD variable in the primary analyses, as we expected that a more advanced LDD change (such as DHN) would associate more strongly with MCs. Because relationships between other LDD features and other MCs may also inform the knowledge base in this area, exploratory analyses were also conducted for the other possible combinations of LDD and MC variables (Table 1).

### Study sample

Participants in LAIDBACK were randomly sampled from primary care, dental, dermatology, and women’s health clinics at the Veterans Affairs (VA) Puget Sound Health Care System, in Seattle, Washington, United States. Sampling was stratified by age so that half of the study sample was <53 years of age.^18^ Participants were excluded for having LBP more than “mildly bothersome” in the last 4 months or a Roland-Morris disability score>3; prior lumbar spine surgery or interventional procedures; serious comorbidities limited expected survival over the 3-year follow-up; impaired cognition or communication; or contraindications to MRI. Briefly, the baseline study sample (n=147) had a mean age of 53 years (range 36-71), 85% were White, 8% were Black/African-American, and 11% were also or solely from other race groups. Consistent with demographics of people with prior United States (US) military service who use the VA system, only 12% were female. Extensive details regarding the study sample characteristics, exclusion criteria and data collected have been previously reported.^18,19^ This research involved deidentified data only that resides at the University of Washington, and did not meet the definition of human subjects research of the University of Washington Human Subjects Division.

### Evaluation of lumbar disc degeneration (LDD) and Modic changes (MCs)

All participants received a standardized lumbar spine MRI protocol through each of the five lumbar interspaces using a Philips 1.5 Tesla MRI system (see below). One of 2 senior neuroradiologists with clinical and research expertise in lumbar spine imaging interpreted the images. The neuroradiologists were aware that participants had no or minimal LBP-related symptoms at baseline but otherwise were blinded to all other participant information. The neuroradiologist recorded the presence or absence of lumbar spine findings at each interspace for the baseline MRIs. Where possible, published definitions or validated grading scales were used for grading lumbar spine MRI findings. A subset of MRIs were interpreted independently by both readers for some MRI findings that were expected to be more prevalent, and reliability was assessed using the unweighted kappa statistic for dichotomous variables and weighted kappa for ordinal variables. MRIs at 3-year follow-up were interpreted using the same methods for reading the baseline MRIs, except that the baseline images and interpretations were available to the neuroradiologists when reading the follow-up images, to minimize variability other than true anatomic changes. As another feature to minimize variability, the same neuroradiologist interpreted the baseline and 3-year follow-up MRIs.

The imaging protocol included the following four imaging sequences :

1. Sagittal T1-weighted, 2-D spin echo, TR/TE 5 600/20, 2 repetitions, 205 3 256 matrix, 26-cm FOV, 4-mm slice thickness, 1-mm skip (scan time, 3 minutes, 30 seconds)
2. Sagittal T2-weighted 2-D fast spin echo, TR/TE 5 3000/85, 2 repetitions, echo train length (ETL) 8, 256 3 256 matrix, 24 3 18 cm FOV, 5-mm slice thickness, 1.5-mm skip (scan time, 3 minutes, 50seconds)
3. Axial T1-weighted, 2-D spin echo, TR/TE 5 700/20, 2 repetitions, 1793 256 matrix, 21-cm FOV, 4-mm slice thickness, 1-mm skip (scan time, 3 minutes, 30 seconds)
4. Axial 2-D fast spin echo, TR/TE 5 4000/72, 2 repetitions, ETL 8, 179 3256 matrix, 16/16 cm fov, 4-mm slice thickness, 1-mm skip (scan time,64 seconds)

Of note, conventional T2-weighted sequences without fat saturation were obtained and not short tau inversion recovery (STIR).

DHN was defined as a decrease in disc height compared with the expected height of a hydrated disc at the same level (a binary variable).^18^ For grading interspaces at L4-L5 and superior, the disc height of the interspace immediately above was used in deciding whether an interspace exhibited DHN. If the disc space above the interspace of interest appeared to be narrowed, and at the L5-S1 level where normal disc height can be more variable, the neuroradiologist’s clinical judgement was exercised. DSI was graded as “mild” if there was a mild diffuse decrease in signal, or a focal moderate or severe decrease involving < 1/3 of the disc area on sagittal images; “moderate’ if there was a moderate diffuse decrease in signal or a focal moderate or severe decrease involving 1/3-2/3 of the disc area ; and “severe” if there was a severe diffuse decrease (no residual high T2 signal) involving >2/3 of the disc area.^18^ Interrater reliability was moderate for DHN (k=0.56) and almost perfect for DSI (weighted k=0.84).

Type I MC was defined as an MC with low T1 and high T2 signal intensity. Type II MC was defined as an MC with high T1 and low to intermediate T2 signal intensity. A type III MC was defined as an MC with low T1 and low T2 signal intensity.^18^ Mixed MC types were not evaluated. Reliability was not evaluated for MCs due to their expected low prevalence.

### Statistical Analysis

In the primary analysis for the first study aim, we conducted a cross-sectional *interspace-level, common-relationship analysis* of the association between DHN and T2MC at baseline, and at 3-year follow-up, using generalized estimating equations (GEE). We adjusted for age, assigned sex at birth, interspace, and MRI time point (baseline vs. 3-year follow-up), and calculated multivariable-adjusted odds ratios (aORs) and 95% confidence intervals (CIs) for the association of DHN with T2MC, using a p-value threshold <0.05 to determine statistical significance. A simplified form of this GEE model is: T2MC ∼ interspace + MRI time point + DHN + sex + age. The DHN variable was the presence vs. absence of DHN at a specific interspace. Similarly, the T2MC variable was the presence vs. absence of T2MC at a specific interspace. A working independence correlation structure was used, for reasons we have described in detail elsewhere.^21^ The L1-L2 interspace (which we refer to simply as the “L1 interspace” for brevity) was chosen as the reference interspace. Importantly, while this was a cross-sectional analysis in which imaging findings at one time point were compared to imaging findings at the *same* time point, it used data from both the baseline and follow-up time point, accounting for time point using GEE. Longitudinal analyses of incident changes were not included in this manuscript due to the previously-reported low incidence of new MRI findings over 3 years in this cohort,^19^ which was expected to provide limited statistical power for analyses of incident changes. We decided *a priori* not to adjust for other variables that were not fixed (e.g., body mass index), as these variables could be consequences of spinal degeneration— including but not limited to the pathway of low back pain and its consequences— and could induce false associations due to collider bias. Collider bias is a type of bias that occurs when an exposure and outcome (or factors affecting these) each cause a common third variable which is conditioned on.^22^ The full model and results for all variables included in the model are provided in Supplemental Table S1.

To address the second study aim, we also calculated aORs and 95% CIs for the association of DHN with T2MC in *interspace-level, interspace-specific analyses*, using a model that was identical to the GEE model from the interspace-level, common-relationship analysis, but also including interaction terms for each interspace with DHN. A simplified form of this GEE model is: T2MC ∼ interspace + MRI time point+ DHN + DHN*interspace + sex + age. In other words, this model adjusted for age, sex, interspace, and MRI time point, as well as interaction terms for DHN x L2 interspace, DHN x L3 interspace, DHN x L4 interspace, and DHN x L5 interspace. The interspace-level, interspace-specific analysis approach is a more specific instance of the interspace-level, common relationship approach that includes these interaction terms, which may allow more detailed investigations of interactions between different spine findings and interspaces, at the potential cost of greater complexity and less statistical precision. We then calculated aORs and 95% CIs for the association of DHN with T2MC in *person-level analyses* adjusting for age, sex, and time point using GEE. A simplified form of this GEE model is: T2MC ∼ interspace + MRI time point+ DHN + DHN*interspace + sex + age. Analogous to the individual-level analyses, the DHN variable was the presence vs. absence of DHN at any lumbar interspace. Similarly, the T2MC variable was the presence vs. absence of T2MC at any lumbar interspace. We then descriptively compared aORs and 95% CIs from interspace-level, common-relationship vs. interspace-level, interspace-specific vs. person-level analyses, expecting to see wider confidence intervals and smaller-magnitude associations in the person-level analysis because this type of summarization essentially introduces a form of measurement error. We used a 4 degree-of-freedom omnibus test to determine if there was a significant difference between the interspace-level, interspace-specific model (a more complex model) interspace-level, common-relationship analyses (a simpler and more general model). This test examines whether interaction terms for DHN x L2 interspace, DHN x L3 interspace, DHN x L4 interspace, and DHN x L5 interspace are significantly different than zero. Our expectation was that if there was no significant difference between the models, the simpler model (the interspace-level, common-relationship model) would be preferred, and differences between interspace-specific associations vs. the overall association produced by the common-relationship model were more likely due to natural variability rather than reflecting real differences between interspaces. We did not conduct power calculations as the sample size was fixed, given well-known concerns regarding *post hoc* power analysis.^23^ The full models and results for all variables included in the model are provided in the Supplemental Tables S2-S4.

We then repeated the analytic steps above in exploratory analyses for the other 7 possible combinations of LDD independent variables (DHN and DSI) and MC dependent variables (T1MC, T2MC, and T3MC), as summarized in Table 1. Although DSI is an ordinal variable, it was analyzed as a continuous variable, as is common practice in analyses of spinal degeneration. Interspace-level analyses used DSI at a specific interspace as a continuous variable. Person-level analyses used the average DSI across all interspaces as a continuous variable. The full models and results for all variables included in the model are provided in the Supplemental Tables S1-S4.

## RESULTS

Of 147 participants, 57% had DHN at ≥1 lumbar interspaces and 95% had at least mild DSI at ≥1 interspaces (Table 2). The prevalence of DHN and severe DSI was generally higher at the L4-S1 interspaces than the L1-L4 interspaces. Type II MCs were present at ≥1 interspaces in 27% of participants, with the lowest prevalence at the L1-L3 interspaces (4-7%) and the highest at L4-S1 (12-14%).

In the primary *interspace-level, common-relationship analysis*, DHN was strongly associated with type II MC (aOR=6.5 [95% CI 3.3-13], p= 4.0 × 10^-8^; Supplemental Table S1). Associations between DHN and type II MC in *interspace-level, interspace-specific* analyses were largest at the L1-2 and L5-S1 interspaces (aOR=13 [95% CI 2.5-73] and aOR=15 [95% CI 4.1-56], respectively), and relatively smaller at the L2-L5 interspaces (aOR at L2-3=2.6 [95% CI 0.44-16], L3-4=4.0 [95% CI 1.1-15], and L4-5=3.8 [95% CI 1.2-16], respectively) (Supplemental Tables S2-3). Associations between DHN and type II MC in *person-level* analyses were smaller (aOR=2.9 [95% CI 1.2-7.2]; Supplemental Table S4).

**Figure 1** summarizes the contrasts between DHN-type II MC associations with the 3 types of analyses, using a logarithmic scale appropriate for odds ratios.^24^ It shows that the DHN-type II MC association point estimate is larger and 95% CI narrower in the interspace-level, common-relationship analysis (shown at the top of the Forest plot) as compared to the person-level analysis (shown at the bottom of the plot). The magnitude of DHN-type II MC association point estimates from interspace-level, interspace-specific analyses are similar to those from the person-level analysis (L2-L5 levels) or larger (L1-2 and L5-S1 levels), but all 95% CIs are substantially wider. When comparing interspace-level, common-relationship vs. interspace-level, interspace-specific analyses as part of the second aim, there was no significant difference between the two (interaction p=0.53, Supplemental Table S2). This indicates that, in this context, the interspace-level, interspace-specific DHN-type II MC associations are not significantly different than the interspace-level, common-relationship DHN-type II MC association (aOR=6.5), with no advantage to the more complex interspace-level, interspace-specific analysis. In both the interspace-level, common-relationship and interspace-level, interspace-specific analyses, no individual interspace (e.g., L5-S1) was significantly associated with type II MC once other factors such as DHN, age, and sex had been accounted for (Supplemental Tables S1, S2, and S4).

**Figure 1.**
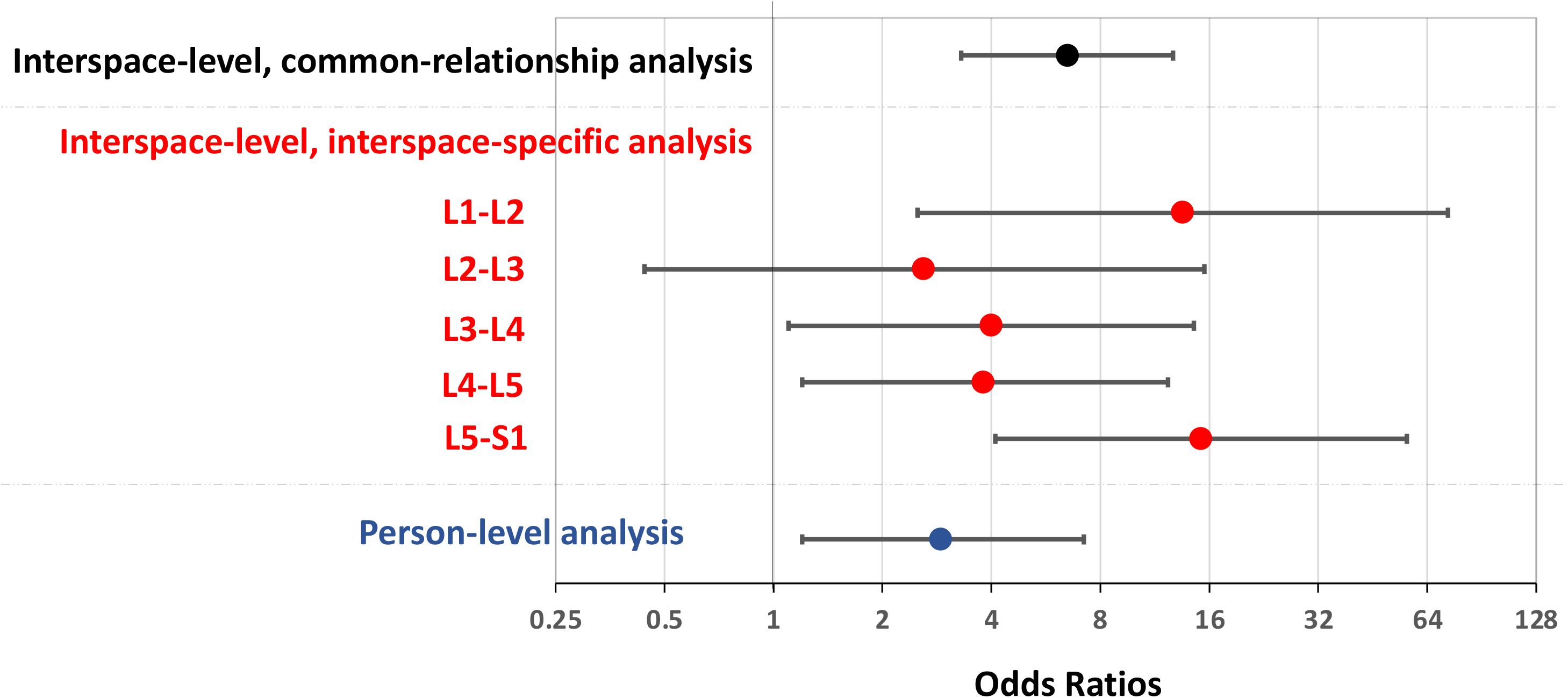
Odds ratios for the DHN-type II MC association. DHN=disc height narrowing, MC=Modic change

The results of exploratory cross-sectional analyses of other predictor-outcome associations including LDD and MC variables are provided in Supplemental Tables S1, S2, and S4 . Of the 7 models for different predictor-outcome relationships, 4 ran without errors in interspace-level, common-relationship analyses, all of which showed significant LDD-MC associations, with the largest being the association of DHN with type I MC (aOR=10, 95% CI 2.5-44]) and the smallest being the association of DSI with type II MC (aOR=2.4, 95% CI [1.7-3.6]). Three models ran without errors in person-level analyses, of which there were two significant LDD-MC associations: the association of DHN with any MC: (aOR=4.2, 95% CI 1.6-11]) and the association of DSI with any MC: (aOR=1.8, 95% CI 1.0-3.4]). Three models ran without errors in interspace-level, interspace-specific analyses. There was a consistent pattern across analyses of larger-magnitude LDD-MC associations with narrower confidence intervals in interspace-level, common-relationship analyses as compared to person-level analyses and interspace-level, interspace-specific analyses (Supplemental Tables S1-S4), when taking into account that for ORs a logarithmic scale is appropriate (rather than a linear scale).^24^ When comparing interspace-level, common-relationship vs. interspace-level, interspace-specific analyses for each LDD-MC relationship, there was no significant difference in any analysis (Supplemental Table S2; interaction p>0.05). As in the primary analysis, no individual interspace was significantly associated with any MC once other factors such as LDD, age, and sex had been accounted for (Supplemental Tables S1, S2, S4).

## DISCUSSION

This study found that DHN and type II MC were strongly associated in an interspace-level, common-relationship analysis, with a larger-magnitude and more precise association than found in a person-level analysis (Figure 1). The same pattern was seen across all exploratory analyses (Supplemental Figures S1-3). These findings illustrate how, in some contexts, interspace-level, common-relationship analyses may yield larger and more precise associations than person-level analyses. While the magnitude of DHN-type II MC associations from interspace-level, interspace-specific analyses differed by interspace in the current study, the absence of statistically significant differences across interspaces suggest that these differences may reflect random variability and did not indicate an advantage to interspace-level, interspace-specific analyses in this context.

To our knowledge this is the first study to comprehensively compare the results of interspace-level and person-level analyses of spinal degeneration findings. Our finding of larger-magnitude associations with interspace-level, common-relationship analyses than person-level analyses within the same study comport with isolated findings scattered across prior reports. A previous cross-sectional, interspace-level, common relationship analysis found a large-magnitude association of any MC with LDD (aOR=6.2, p<0.001).^16^ This contrasts with much smaller-magnitude any MC-LDD associations in a person-level analysis from another study (OR for DHN=1.3 and OR for DSI=1.2, both p-values <0.001).^3^ The current findings indicate that such between-study differences in effect size are more likely due to the different analytic methods involved, and less likely explained by differences in study populations or designs.

Interspace-level analyses may also have other advantages. In contrast to person-level analyses, interspace-level analyses allow evaluation of the relationship between an interspace and a specific lumbar degeneration finding that is independent of lumbar degeneration findings at the same interspace. An example of this is the current study’s finding that spinal interspace was not significantly associated with MCs once the LDD variable at the same interspace was adjusted for. This is notable, as most prior studies find differences in MC prevalence by interspace, with lower prevalences at L1-L4, and higher prevalences at L4-S1.^5,16,25^ These findings taken together with points from preceding paragraphs suggest the possibility that spinal interspace is not associated with MCs once LDD at the same level has been accounted for because LDD at an interspace may largely explain the subsequent development of an MC at the same interspace. This type of inference is permitted by interspace-level analytic approaches but would be obscured by person-level analyses.

On the other hand, the use of interspace-level analyses shifts interpretation to the spinal level rather than the person. Interspace-level analyses may be more appropriate for certain research contexts, such as when examining a biological relationship that credibly has its predominant effect at the same interspace (and <u>not</u> an equal effect at all lumbar interspaces). For example, when conducting a study of experimental disc injury, a researcher may believe that disc injury would be more likely to cause early disc degeneration at the involved interspace, rather than simultaneously causing degeneration at multiple lumbar interspaces. In this situation, an interspace-level analysis is probably more appropriate than a person-level analysis. If, however, a research study was examining the possible effect of cigarette smoking on disc desiccation, and a researcher expects *a priori* that smoking causes disc desiccation uniformly at multiple lumbar interspaces, a person-level analytic approach may be more suitable than an interspace-level analysis. Researchers should consider interspace-level analyses as potential alternatives or complements to person-level analyses, depending on the specific research question in a given study.

A study strength is that we publicly posted our analysis plan *a priori*, to prevent data mining.^20^ While the study’s abundant statistical power for evaluating the DHN-type II MC relationship in an interspace-level, common-relationship analysis is shown by the highly significant association found (p=4.0 × 10^-8^), a potential limitation is that the study may have been underpowered when comparing interspace-level, interspace-specific vs. interspace-level, common-relationship models. Given the conceptual advantages of interspace-level, interspace-specific models in emulating the complexity of the lumbar spine, in which biomechanics, disc nutrition, and other factors are thought to affect different interspaces differently, we suggest that future studies use such models in the larger cohorts now available.^1,6,26^

In summary, this study illustrates how interspace-level analytic approaches may offer certain advantages to person-level analyses that can be useful in understanding relationships between spinal degeneration findings.

## Data Availability

The study original informed consent processes and institutional policies do not allow external individual-level data sharing. The analytic code for this work will be made available upon request.

