## Supplementary material for "Advantages to interspace-level analyses over person-level analyses when studying relationships between lumbar spinal degeneration findings : a cross-sectional study": Tables

| **Table 1. Overview of the study aims, designs, primary analyses, and exploratory analyses.** | |
| --- | --- |
| **Aim 1: To examine associations between LDD and type II MCs using an *interspace-level, common-relationship* analytic approach.** *Hypothesis: LDD and type II MC are significantly associated when using this approach.* | |
| **Aim 2:** **To descriptively compare associations between LDD and type II MC when using (1) an *interspace-level, common-relationship* approach, (2) an *interspace-level, interspace-specific* approach, and (3) a *person-level* approach*.*** *Hypothesis: A model using an interspace-level, interspace-specific approach is significantly different from a model using an interspace-level, common-relationship approach.* | |
| **Primary analysis^a^** | |
| **Independent variable** | **Dependent variable** |
| Disc height narrowing (DHN) | Type II Modic change (MC) |
| **Exploratory analyses** | |
| **Independent variable(s)** | **Dependent variable(s)** |
| DHN, disc signal intensity (DSI) | Type I MC, type II MC, Type III MC, any MC |
| ^a^Cross-sectional analyses of DHN and type II MC using generalized estimating equations (GEEs), adjusting for age, sex, interspace, and time point. One statistical comparison was made for each of the 2 Aims, each using a p-value significance threshold of 0.05.  ^b^An analogous approach to the primary analysis was taken for the exploratory analyses, which included 2 predictor variables and 4 outcome variables, for a total of 7 exploratory statistical comparisons (not including the association of DHN with MC, which was the primary analysis) | |

| **Table 2. Prevalence of lumbar disc degeneration (LDD) and Modic changes (MC)*** | | | | | | |
| --- | --- | --- | --- | --- | --- | --- |
| **Prevalence at baseline (n=147).** | | | | | | |
| **LDD at baseline** | | | | | | |
|  | **L1** | **L2** | **L3** | **L4** | **L5** | **Any interspace** |
| DHN | 20 (14%) | 17 (12%) | 23 (16%) | 41 (28%) | 45 (31%) | 84 (57%) |
| DSI |  |  |  |  |  |  |
| None | 60 (41%) | 56 (38%) | 46 (31%) | 33 (22%) | 35 (24%) | 8 (5%) |
| Mild | 44 (30%) | 37 (25%) | 32 (22%) | 37 (20%) | 19 (13%) | 92 (63%) |
| Moderate | 24 (16%) | 36 (24%) | 35 (24%) | 36 (24%) | 34 (23%) | 99 (67%) |
| Severe | 19 (13%) | 18 (12%) | 34 (23%) | 41 (28%) | 59 (40%) | 83 (56%) |
| Any LDD | 87 (59%) | 91 (62%) | 101 (69%) | 114 (78%) | 112 (76%) | 139 (95%) |
| **Modic changes at baseline** | | | | | | |
|  | **L1** | **L2** | **L3** | **L4** | **L5** | **Any interspace** |
| Type I MC | 1 (1%) | 1 (1%) | 1 (1%) | 0 | 2 (1%) | 5 (3%) |
| Type II MC | 7 (5%) | 6 (4%) | 11 (7%) | 17 (12%) | 20 (14%) | 39 (27%) |
| Type III MC | 0 | 1 (1%) | 0 | 0 | 1 (1%) | 2 (1%) |
| Any MC | 8 (5%) | 8 (5%) | 12 (8%) | 17 (12%) | 23 (16%) | 43 (29%) |
| *There was no missing data | | | | | | |
