## Supplemental Figures for "Advantages to interspace-level analyses over person-level analyses when studying relationships between lumbar spinal degeneration findings : a cross-sectional study"

**Figure S1. Odds ratios for the DHN-any MC association**

DHN=disc height narrowing, MC=Modic change

**Figure S2. Odds ratios for the DSI-type II MC association**

DSI=disc signal intensity loss, MC=Modic change

**Figure S3. Odds ratios for the DSI-any MC association**

DSI=disc signal intensity loss, MC=Modic change
